# Interaction of mitochondrial polygenic score and environmental factors in LRRK2 p.Gly2019Ser parkinsonism

**DOI:** 10.1101/2023.01.02.23284113

**Authors:** Theresa Lüth, Carolin Gabbert, Inke R. König, Amke Caliebe, Sebastian Koch, Björn-Hergen Laabs, Faycel Hentati, Samia Ben Sassi, Rim Amouri, Malte Spielmann, Christine Klein, Anne Grünewald, Matthew J. Farrer, Joanne Trinh

## Abstract

The objective of our study was to investigate the impact of the mitochondrial polygenic score (MGS) and lifestyle/environmental data on age at onset in LRRK2 p.Gly2019Ser parkinsonism (*LRRK2*-PD) and idiopathic Parkinson’s disease (iPD).

In this study, we included N=486 patients with *LRRK2*-PD and N=9259 patients with iPD from AMP-PD, Fox Insight, and a Tunisian Arab-Berber founder population. Genotyping data was utilized to perform the MGS analysis, using 14 Single Nucleotide Polymorphisms (SNPs) from genes causally associated with mitochondrial function and PD risk. Additionally, lifestyle and environmental data were obtained from the PD risk factor questionnaire (PD-RFQ). Correlation analyses and linear regression models were used to assess the relationship between MGS, lifestyle/environment, and AAO.

We observed that higher MGS was associated with earlier AAO in patients with *LRRK2*-PD (p=4.0×10^−4^, β=-0.18) but not in patients with iPD. A correlation between MGS and AAO was visibly stronger in European ancestry *LRRK2*-PD patients (p=0.01, r=-0.16) than in Tunisian Arab-Berber patients (p=0.44, r=-0.05). We found that the MGS interacted with coffee (p=0.03, β=-0.38) and caffeinated soda consumption (p=0.03, β=-0.37) in *LRRK2*-PD and with caffeine soda consumption (p=0.047, β=-0.22) and pesticide exposure (p=0.02, β=-0.37) in iPD. Thus, patients with a high MGS had an earlier AAO only if they consumed caffeine or were exposed to pesticides.

The MGS related to mitochondrial function was associated with AAO in *LRRK2*-PD but not iPD with an ethnic-specific effect. Caffeine consumption or pesticide exposure interacted with MGS to predict PD AAO. Our study suggests gene-environment interactions as modifiers of AAO in *LRRK2*-PD.

## Introduction

The age at onset (AAO) of Parkinson’s disease (PD), as well as the risk of developing the disease, are known to be affected by genetic and environmental factors.^1-5^ In terms of genetics, monogenic forms and strong risk factors account for ∼10% of PD cases.^6^ Among these cases, the most common monogenic cause is the Leucine-rich repeat kinase 2 (LRRK2) p.Gly2019Ser mutation. Besides monogenic forms and other genetic variants, PD can be explained by the interplay of complex genetics and lifestyle or environmental factors. One way to assess the cumulative effect of genetic variants on disease risk or AAO is by deriving and using a polygenic score (PGS).^7,8^ Previously, a mitochondrial polygenic score (MGS) was derived and composed of genes involved in mitophagy, mitochondrial bioenergetics and proteostasis pathways. The study nominated 14 SNPs as causally associated with mitochondrial function and PD risk by Mendelian randomization.^9^ Biologically, mitochondria are essential key players in PD pathogenesis. In particular, respiratory chain, mitophagy, and mitochondrial biogenesis impairment are associated with PD.^10^

LRRK2 localizes to the cytosol as well as to the mitochondria in the cells. Additionally, fibroblasts derived from patients with *LRRK2*-PD showed reduced NADH dehydrogenase activity and increased mitochondrial mass, mtDNA copy number and nuclear factor erythroid 2-related factor 2 (Nrf2) expression.^11^ In macrophages, the LRRK2 p.Gly2019Ser mutation interferes with mitochondrial homeostasis and alters cell death pathways.^12^ A recent study reported that the seeding of A53T alpha-synuclein oligomerization happens especially at mitochondrial membranes in neurons, which can lead to respiratory chain impairments and a subsequent increase in reactive oxygen species (ROS).^13^ As alpha-synuclein pathology is an important hallmark in *LRRK2*-PD and iPD, this is another molecular link between PD pathogenesis and mitochondrial impairment.

PD susceptibility has consistently been associated with lifestyle and environmental factors. Several meta-analyses have highlighted the negative association between smoking and PD risk.^14^ It has been demonstrated that smoking status correlates with later AAO in iPD.^1,15,16^ Additionally, caffeine and non-steroidal anti-inflammatory drug (NSAIDs) consumption was associated with reduced iPD risk and later AAO.^1,14^ Analogous to iPD, smoking and caffeine consumption are associated with later onset in *LRRK2*-PD.^5,17^ In a study including affected and unaffected *LRRK2* mutation carriers, NSAIDs (i.e., aspirin and ibuprofen) users had reduced odds of developing PD.^18^ In addition to lifestyle and environment, genetic modifiers of the AAO have been identified as well. There is evidence that variants in the *DNM3*^*19*^ and *CORO1C*^*20*^ genes are associated with AAO in *LRRK2*-PD.

The interaction of mitochondrial-related genes, lifestyle and environment has not been thoroughly investigated. Importantly, mitochondria are at the interface of environmental impacts in the cell. For example, pesticide exposure can produce reactive oxygen species (ROS) by redox cycling, inducing mtDNA damage.^10^ In addition, smoking and vaping have been shown to be associated with mitochondrial gene dysregulation^21^ and there is evidence that caffeine affects mitochondrial bioenergetics^22^ and increases mitochondrial function.^23^ Our study aimed to analyze these interactions between the MGS and lifestyle and environment in *LRRK2*-PD and iPD.

## Materials and methods

### Study demographics, genetics and environmental data

Three datasets with genetic, environmental and lifestyle data were included in this study: AMP-PD, Fox Insight, and a cohort from the Tunisian Arab-Berber population. In total, 9745 patients were included in our study: 486 patients with *LRRK2*-PD (AMP-PD: 127, Fox Insight: 154, Tunisian cohort: 205) and 9259 patients with iPD (AMP-PD: 2077, Fox Insight: 6949, Tunisian cohort: 233). For patients with *LRRK2*-PD, the mean AAO was 58.2 years (SD=11.1) and the mean age at examination (AAE) was 66.7 years (SD=12.4). The mean AAO of patients with iPD was 61.2 years (SD=10.2) and the mean AAE was 65.2 years (SD=9.6) (Table 1).

**Table 1.**
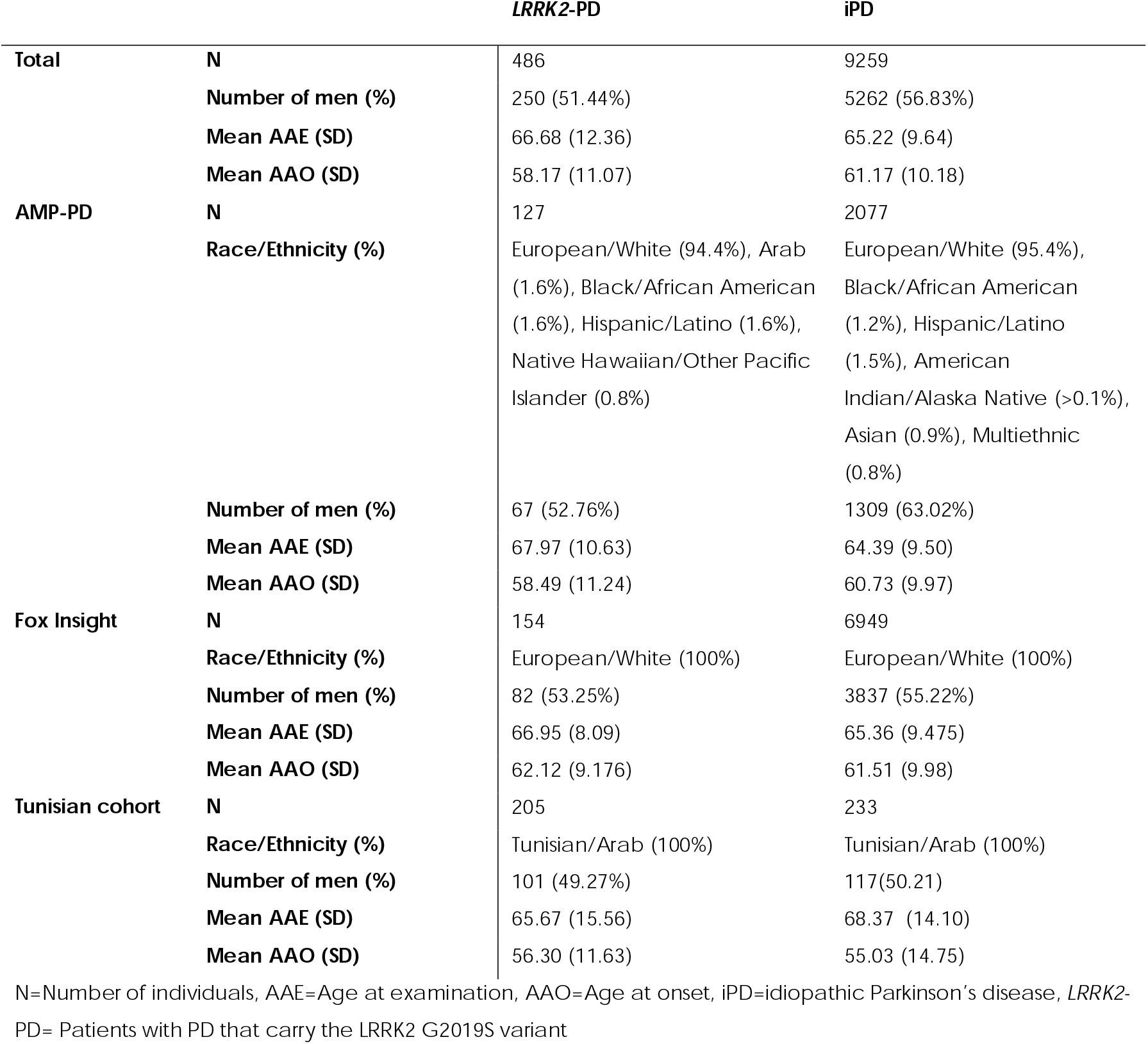
Demographics of patients with PD from the three investigated data sets

AMP-PD contains whole-genome sequencing (WGS) data from four harmonized cohorts. All samples of the AMP-PD dataset were processed by the TOPMed Freeze 9 Variant Calling Pipeline for joint genotyping.^24^ The majority of the patients of the AMP-PD cohort were of European descent (∼95%) and the remaining ∼5% were of Arab, African American, Hispanic, Asian, Native Hawaiian, or Alaskan descent, with self-reported ethnicity/race. The Fox Insight dataset is a cohort within the Michael J. Fox Foundation (MJFF).^25^ For Fox Insight, the genetic data (array-based genotyping) of the patients with PD were provided by 23andMe, as previously described.^25^ Three platforms were used to perform the genotyping within the Fox Insight data: V3 (Illumina OmniExpress□+□BeadChip), V4 (fully custom array) and V5 (customized Illumina Infinium Global Screening Array).^25^ All patients with PD included from the Fox Insight cohort in this study were of self-reported European ancestry. Lastly, we included a cohort recruited from the Tunisian Arab-Berber population, where the genotyping data was obtained from Affymetrix and Illumina MEGA arrays, as previously described.^19^

In the Fox Insight and Tunisian cohort, lifestyle and environmental information were assessed with the PD Risk Factor Questionnaire (PD-RFQ) for tobacco use, caffeine consumption, and pesticide exposure.^26^ However, the AMP-PD dataset did not assess environment and lifestyle data with the PD-RFQ. Therefore, available environmental/lifestyle data in the AMP-PD cohort was not used to maintain consistency and utilized the more detailed data of the Fox Insight and Tunisian cohorts.

According to the PD-RFQ, tobacco use is defined as smoking at least one cigarette per day for more than six months, smoking more than 100 cigarettes in your lifetime, or using smokeless tobacco at least once per day for more than six months. Caffeine consumption means drinking coffee, black tea, green tea or caffeinated soda at least once per week for more than six months. The duration was defined as the number of years the caffeinated beverage was consumed until the AAO. The dosage of a caffeinated beverage is defined as the number of cups per week. We set the dosage to zero cups per week for patients who did not consume a particular caffeinated beverage. In addition, we also included the cumulative dosage of caffeine using the number of cups consumed for individual drinks added up. Lastly, pesticide exposure is exposure to any kind of pesticide in work or non-work settings ever in your lifetime. The exposure duration is estimated as the number of years exposed to pesticides until the AAO.

### Mitochondrial polygenic score analysis

The genetic datasets from AMP-PD, Fox Insight, and the Tunisian cohort were stored in a binary PLINK format^27^. The same quality control filtering steps were applied to all three datasets (minor allele frequency >0.01, missingness per sample <0.02, missingness per SNP<0.05 and Hardy-Weinberg equilibrium >1×10^−50^) using PLINK v1.9. The Fox Insight dataset was imputed using the Michigan Imputation Server^28^ in combination with the Haplotype Reference Consortium v1.1 reference panel.^29^ As the Tunisian dataset is of North African background, we performed the imputation on our in-house computer cluster, using SHAPEIT^30^ and IMPUTE2^31^ in combination with the 1000 Genomes Project Phase 3.^32^ Genotyping data for AMP-PD was obtained from WGS.

The MGS was calculated using the PLINK score function, based on the 14 SNPs and corresponding weights published by Billingsley *et al*..^9^ In order to harmonize the MGS between cohorts, we only used SNPs that were consistently present in all three datasets. Therefore, 9/14 SNPs were used to calculate the MGS across all three cohorts (rs9185, rs57668191, rs139439, rs17788127, rs3824783, rs4886636, rs7157678, rs11038689, rs9905991). When separately investigating individual cohorts, 12/14 SNPs could be used for Fox Insight and 14/14 SNPs for AMP-PD. The obtained MGS was standardized to a percentage, ranging from zero to 100, for better interpretation of the effect size in the following statistical analysis, where zero equals the lowest possible MGS and 100 equals the highest possible MGS. The standardized MGS was calculated with this formula: MGS_std_ = (MGS – lowest possible MGS) ÷ (highest possible MGS – lowest possible MGS) × 100.

### Principal component analysis

In order to visualize populations, we used PLINK to perform a principal component analysis (PCA) based on common SNPs (maf > 0.3). As a reference, we used the publicly available 1000 Genomes Project data. After filtering for common SNPs, we excluded variants not present in all four data sets and then the PCA was performed. Finally, the output was visualized with R and colored according to the study cohort and race/ethnicity.

### Statistical analysis

Statistical analyses were performed with Graphpad Prism v9.4.0 and R v4.0.3.^33,34^ The analyses in this study were exploratory and p-values were not corrected for multiple testing. The association between AAO and MGS was first assessed using multiple regression analyses. Then, multiple linear regression models were also used to evaluate the interaction between MGS and environmental and lifestyle factors.

In our linear regression models, we used AAO as a dependent variable and the standardized MGS (MGS_std_) as an independent variable. We included sex, study cohort (i.e., AMP-PD, Fox Insight, or Tunisian cohort), and ethnicity/race (i.e., European/White, Tunisian/Arab, and others) in the regression models to adjust for potential confounders. We summarized all ethnicities/races besides European/White and Tunisian/Arab to “others” due to the low abundance in our datasets.

Lifestyle and environmental exposure were set as dichotomous independent variables (yes/no) in our linear regression models. Here, we did not include ethnicity/race as a covariate, as only Fox Insight and the Tunisian cohorts were used. All patients from the Fox Insight dataset were of European/White ancestry and all patients from the Tunisian dataset were of Tunisian/Arab ancestry. In additional analyses, caffeine consumption dosage (number of cups), caffeine consumption duration (number of years until AAO) and pesticide exposure duration (number of years until AAO) were modelled as continuous independent variables. We further performed Kaplan-Meier analyses to assess the difference in AAO of patients with high or low MGS_std_ and pairwise comparison was performed using the log-rank test. For the stratification, we defined “high MGS_std_” as higher or equal to the median MGS_std_ and “low MGS_std_^“^ was defined as lower than the median MGS_std_.

### Data availability

Data sharing is not applicable to this article as no new data were created or analysed in this study. Data used in the preparation of this manuscript were obtained from the Fox Insight database (https://foxinsight-info.michaeljfox.org/insight/explore/insight.jsp) on 18/10/2020. For up-to-date information on the study, visit https://foxinsight-info.michaeljfox.org/insight/explore/insight.jsp. Data used in the preparation of this article were obtained from the Accelerating Medicine Partnership® (AMP®) Parkinson’s Disease (AMP PD) Knowledge Platform. For up-to-date information on the study, visit https://www.amp-pd.org.

## Results

### Association between MGS and AAO in PD analysis

First, we analyzed the association between the MGS_std_ and the AAO in patients with *LRRK2*-PD. The MGS_std_ was inversely correlated with the AAO (*r*=-0.15, *p*=0.0008, *N*=486, Figure 1A). The higher the MGS_std_, the earlier the AAO in *LRRK2*-PD. We investigated this relationship using linear models and confirmed the negative association (*p*=0.042, β=-0.11, *SE*=0.05, Table 2). Thus, if the MGS_std_ is increased by 1%, the AAO is approximately one month earlier. As the AAO is earlier in females compared to males^35^ and the AAO and MGS_std_ vary between the three cohorts and ethnicities, we included sex, study site and ethnicity as covariates in the regression models (Table 2). Interestingly, when stratifying the data for the two most abundant ethnicities/races (i.e., European/White or Tunisian/Arab) to analyze the MGS and AAO association, a negative correlation in the same magnitude as before was observed for patients of European descent (*r*=-0.16, *p*=0.01, Supplementary Table 1). However, when looking at the patients of Tunisian Arab-Berber descent, the negative correlation is not as pronounced (*r*=-0.05, *p*=0.44). We did not observe a correlation between MGS_sdt_ and AAO in patients with iPD (Figure 1B). Furthermore, we did not detect an association using the regression model.

**Figure 1.**
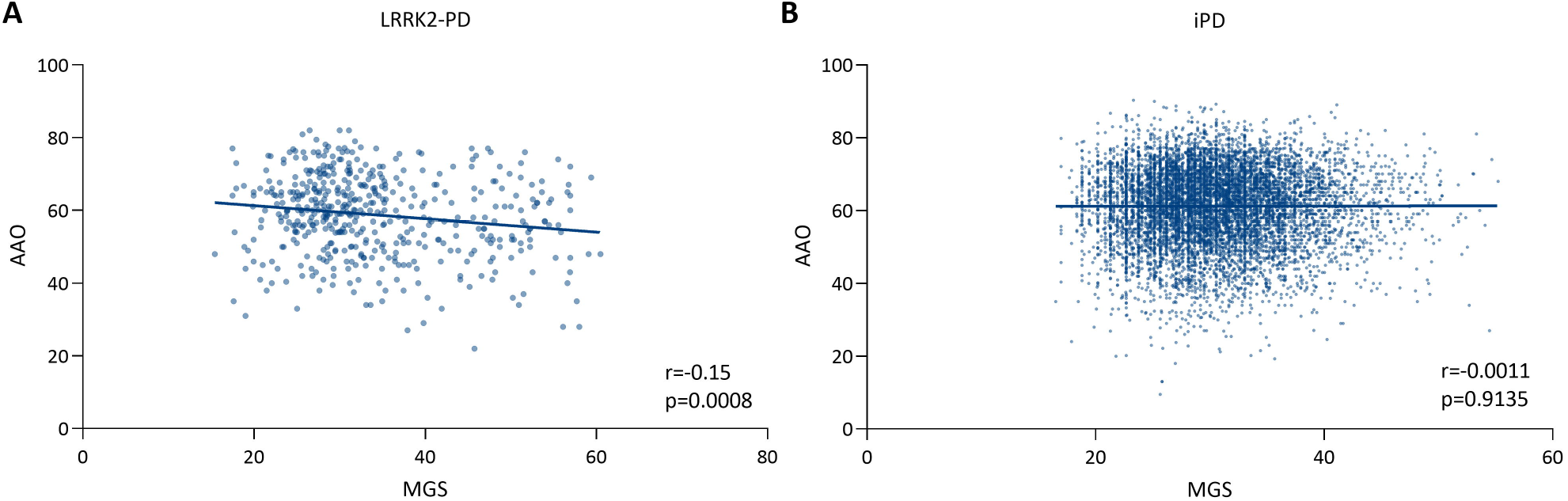
Relationship between age at onset (AAO) and mitochondrial polygenic score (MGS). The correlation plots show the association between MGS and AAO in patients with Parkinson’s disease carrying the LRRK2 p.Gly2019Ser mutation (*LRRK2*-PD) (**A**) or patients with idiopathic PD (iPD) (**B**). r = Spearman’s rank correlation coefficient, p = Spearman’s exploratory p-value.

**Table 2.** Association between the mitochondrial polygenic score (MGS) and the age at onset (AAO) in patients with *LRRK2*-PD and idiopathic PD.

|  | *LRRK2*-PD |  |  | iPD |  |  |
| --- | --- | --- | --- | --- | --- | --- |
|  | Estimate | Std. Error | p-value | Estimate | Std. Error | p-value |
| Association between MGS and AAO with sex, study cohort and race/ethnicity as covariates<sup>1</sup> |  |  |  |  |  |  |
| **MGS<sub>std</sub>** | -0.11 | 0.05 | 0.042* | 0.01 | 0.02 | 0.56 |
| **Sex Male** | 1.83 | 0.98 | 0.06 | 0.91 | 0.21 | 2.48 × 10<sup>-5</sup>* |
| **Study Fox Insight** | 2.62 | 1.33 | 0.049* | 0.87 | 0.28 | 0.002* |
| **Study Tunisian cohort** | 7.14 | 3.66 | 0.05 | -3.56 | 1.23 | 0.004* |
| **Ethnicity Tunisian/Arab** | NA | NA | NA | NA | NA | NA |
| **Ethnicity White** | 9.42 | 3.71 | 0.011* | 2.06 | 1.05 | 0.05 |

iPD=idiopathic Parkinson's disease, *LRRK2*-PD= Patients with PD that carry the *LRRK2* G2019S variant,
MGS<sub>std</sub>=Mitochondrial polygenic score, \* p-value < 0.05
<sup>1</sup>glm(formula = AAO ~ MGS<sub>std</sub> + Sex + Study cohort + Ethnicity, family = gaussian)
Baseline categories: Sex=Female, Study=AMP-PD and Ethnicity=Other (i.e., not white or Tunisian/Arab)

### Effect of lifestyle factors and environmental exposure on AAO

We focused our analysis on known protective (smoking and caffeine consumption) and risk factors (pesticide exposure) in PD using regression models including sex and study cohort as a covariate.

We observed no association of smoking with AAO in *LRRK2*-PD (*p*=0.194, β=2.67, *SE*=2.05, Supplementary Table 2). In iPD, smoking was associated with later AAO (*p*=0.001, β=1.45, *SE*=0.46). To thoroughly assess the relationship between caffeine and AAO in PD, we analyzed coffee, black tea, green tea and caffeinated soda consumption. In *LRRK2*-PD, the only caffeinated beverage associated with later AAO was black tea (*p*=0.002, β=6.10, *SE*=1.97). In patients with iPD, coffee consumption was associated with later AAO (*p*=2.0×10^−4^, β=2.06, *SE*=0.55). Green tea, however, was not associated with AAO in *LRRK2*-PD or iPD. Caffeinated soda was associated with earlier AAO in *LRRK2*-PD (*p*=0.035, β=-4.29, *SE*=2.01) and iPD (*p*=8.8×10^−6^, β=-2.41, *SE*=0.54). Lastly, we did not observe an association between pesticide exposure in a work setting and AAO in *LRRK2*-PD but in iPD (*p*=0.048, β=-1.52, *SE*=0.76, Supplementary Table 3).

### Interactions between lifestyle, environment and MGS on AAO

Next, we explored the interaction of lifestyle/environment and MGS on AAO in a linear regression model (Table 3).

**Table 3.**
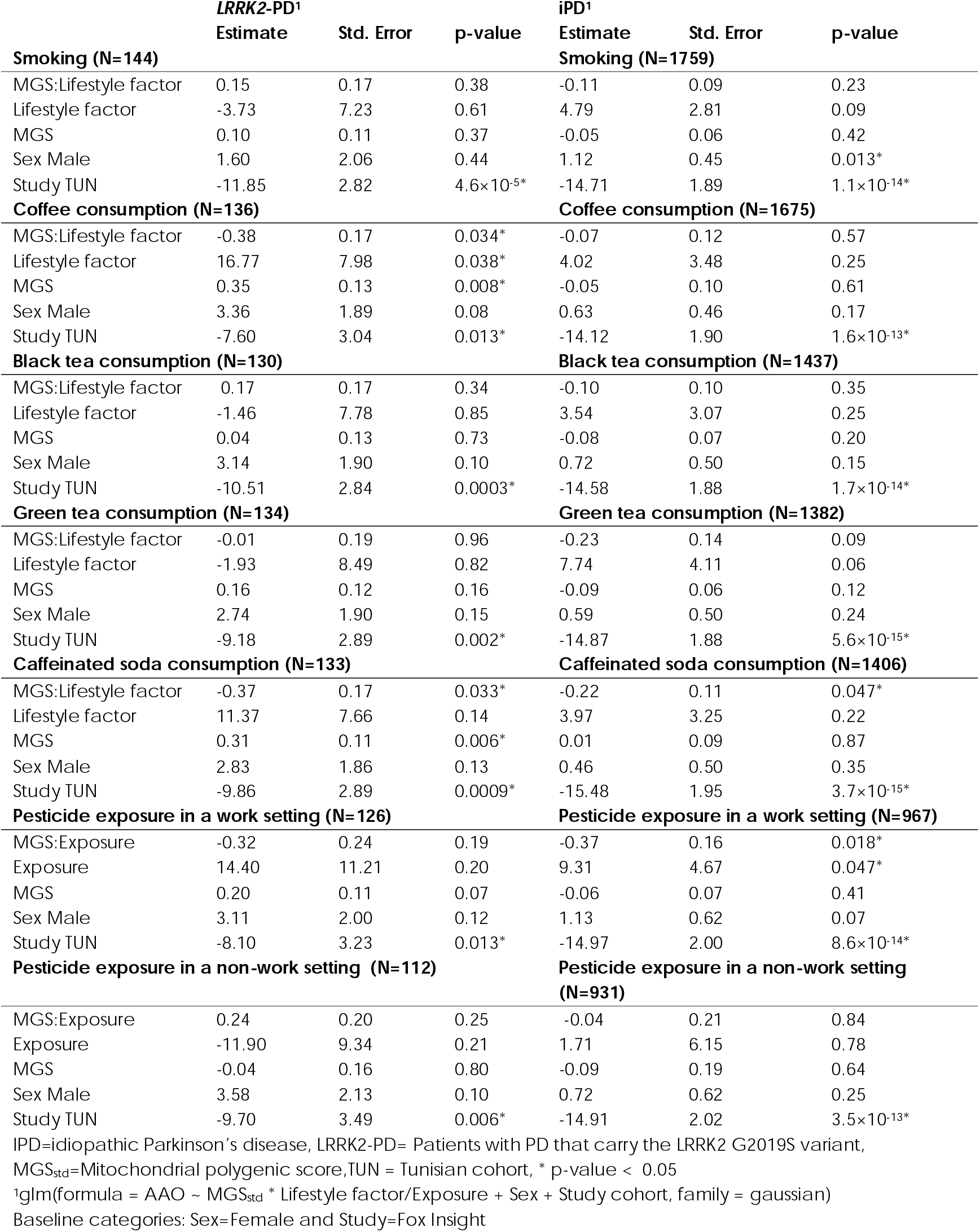
Interaction between the mitochondrial polygenic score, lifestyle factors and pesticide exposure and the age at onset in patients with*LRRK2*-PD and idiopathic PD.

As we did not find a significant interaction between MGS_std_ and smoking in *LRRK2*-PD and iPD, we did not continue to look into the dosage and duration of tobacco use.

Interestingly, we detected an interaction between MGS_std_ and coffee consumption with the AAO of *LRRK2*-PD patients (*p*=0.034, β=-0.38, *SE*=0.17). There was also an interaction between MGS_std_ and caffeinated soda consumption (*p*=0.033, β=-0.37, *SE*=0.17) in *LRRK2*-PD. In other words, our results suggest that the association between the MGS_std_ and AAO is dependent upon coffee or caffeinated soda consumption. To validate this potential interaction, we performed Kaplan-Meier analyses which showed an earlier AAO in patients with a high MGS_std_ who consumed one of the caffeinated beverages (Figure 2) in *LRRK2*-PD. In comparison, the AAO was later in patients with a high MGS_std_ who did not drink coffee or caffeinated soda. The median AAO of patients that consumed coffee and had a high MGS_std_ was 55.5 years, compared to *LRRK2*-PD patients with a low MGS_std_ that consumed coffee at 61.2 years (*p*=0.113, Figure 2). The difference in AAO in patients that consumed caffeinated soda was even more pronounced (high MGS_std_: median AAO=50.0, low MGS_std_: median AAO=61.5, *p*=0.010). Thus, coffee consumers with a high MGS_std_ had a ∼six years earlier median AAO and caffeinated soda consumers with a high MGS had a ∼11 years earlier median AAO than *LRRK2*-PD patients with a low MGS. In comparison, the median AAO was only ∼four years earlier in all *LRRK2*-PD patients, unstratified for any lifestyle factor. Interestingly, the AAO was later in *LRRK2*-PD patients with a high MGS that did not consume these caffeinated beverages.

**Figure 2.**
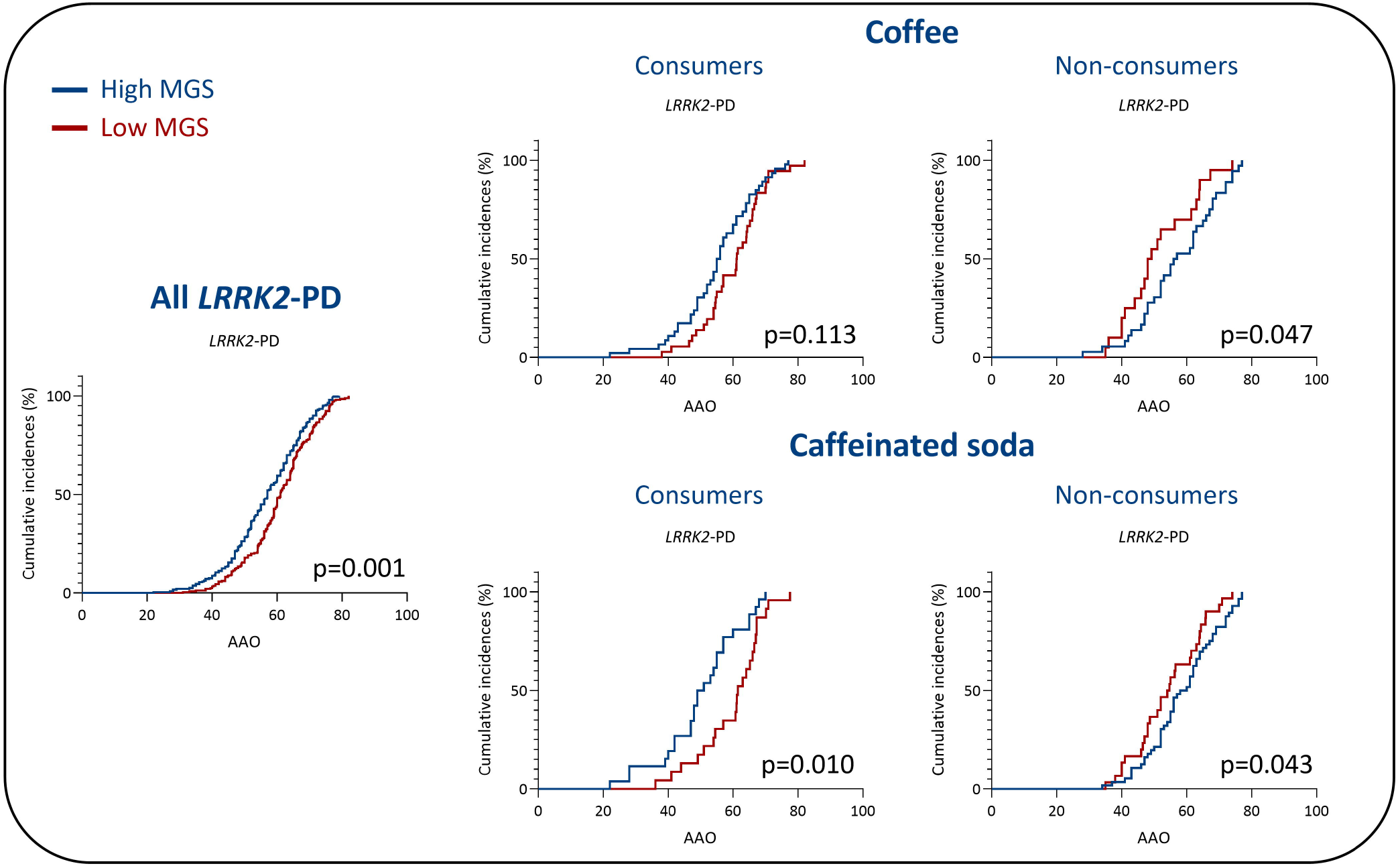
Relationship between age at onset (AAO), mitochondrial polygenic score (MGS) and caffeine consumption. Kaplan-Meier plots show the difference in AAO of patients with *LRRK2*-PD and high MGS or low MGS. The patients were plotted unstratified and stratified by coffee or caffeinated soda consumption. p=Log-rank test p-value

In iPD, an interaction between caffeinated soda consumption and MGS_std_ was also present (*p*=0.047, β=-0.23, *SE*=0.14). There was no interaction between coffee consumption and MGS_std_. The Kaplan-Meier analyses showed an earlier AAO in patients with a high MGS_std_ who consumed caffeinated soda in iPD (Figure 3). The median AAO of patients with higher MGS_std_ was 60.9 years, whereas the median AAO in patients with lower MGS_std_ was 62.7 years (*p*=0.001, Figure 3). There was no difference in AAO of patients with a high or low MGS who did not consume caffeinated soda.

**Figure 3.**
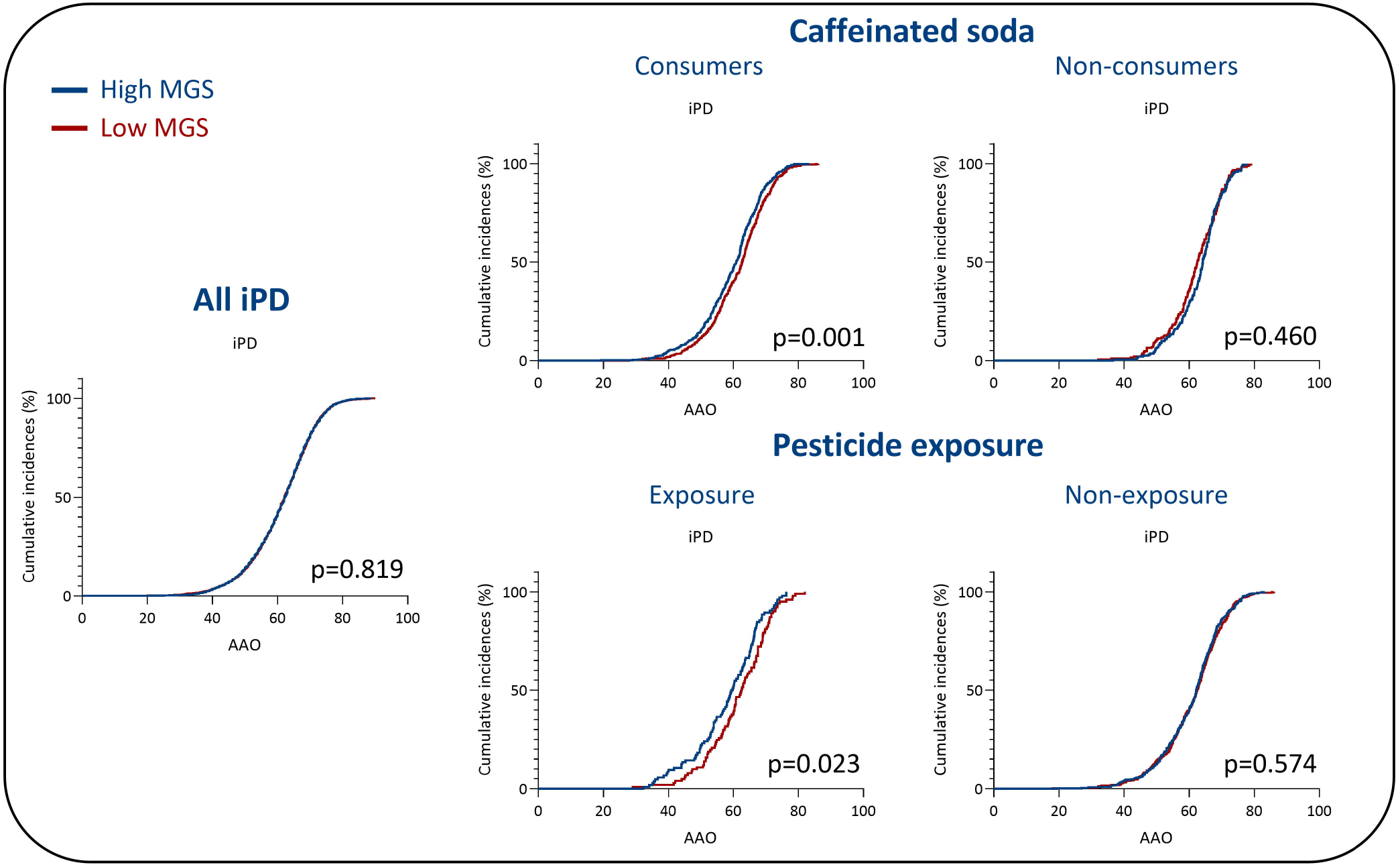
Relationship between age at onset (AAO), mitochondrial polygenic score (MGS) and caffeine consumption or pesticide exposure. Kaplan-Meier plots show the difference in AAO of patients with iPD and high MGS or low MGS. The patients were plotted unstratified and stratified by caffeinated soda consumption or pesticide exposure. p=Log-rank test p-value

For black tea and green tea, we did not observe an interaction with the MGS_std_ (Table 3). We then proceeded to investigate the dosage and duration of caffeinated beverage consumption that showed an interaction with MGS_std_ (i.e., coffee and caffeinated soda, Supplementary Tables 4 and 5). However, we did not detect an interaction in *LRRK2*-PD or iPD. Additionally, we looked into the cumulative dosage of coffee and caffeinated soda together, which also did not show an interaction with MGS_std_.

Lastly, there was no interaction between MGS_std_ and pesticide exposure in a work or non-work setting in *LRRK2*-PD.

In contrast, an interaction between MGS_std_ and pesticide exposure in a work setting was seen in patients with iPD (*p*=0.018, β=-0.37, *SE*=0.16). In the Kaplan-Meier analysis, patients with iPD exposed to pesticides and with a higher MGS_std_ had an earlier AAO than those with a lower MGS_std_ (Figure 3). The median AAO of patients with high MGS_std_ was 59.6 and with low MGS_std_ 62.5 (*p*=0.023). There was no difference in AAO of patients that were not exposed to pesticides. To perform a dose-dependent analysis, we investigated the interaction of MGS_std_ and the duration of pesticide exposure until AAO in iPD. However, there was no interaction between MGS_std_ and exposure duration (Supplementary Table 5).

In order to account for potential biases coming from the different cohorts or ethnicities, we utilized the large sample size of iPD patients in the Fox Insight cohort of European/White descent. We repeated the analysis using the regression models with the iPD patients of Fox Insight only (Supplementary Table 6). There was an interaction between MGS_std_ and caffeinated soda (*p*=0.017, β=-0.26, *SE*=0.11) as well as between MGS_std_ and pesticide exposure at work (*p*=0.012, β=-0.39, *SE*=0.16).

## Discussion

Gene-environment interactions are relevant as onset modifiers of *LRRK2*-PD and iPD. The main strength of this study is the size of the study cohort consisting of three large cohorts. In addition, we utilized the thorough overlap of genetic, lifestyle, and environmental data of two cohorts to comprehensively investigate the relationship between MGS and AAO in PD. We see a robust relationship between the MGS and AAO in *LRRK2*-PD even after adjusting for potentially confounding covariates (i.e., sex, cohort or ethnicity). To our knowledge, we demonstrate a novel association between MGS and later AAO in *LRRK2*-PD. Furthermore, the diverse ethnic background of the patients in this study shows population-specific effects of the MGS. Though we see an overall association between the MGS and AAO, when separating the cohorts, the association was found to be more pronounced in the European cohorts and visibly weaker in the Tunisian/Arab cohort (Supplementary Table 1). It is well known that population- or ethnic-specific background is a key factor in polygenic scores and it is important for future studies to be inclusive of patients from diverse backgrounds.^36-38^ To illustrate the importance of the study population in genetic scores, we performed a principal component analysis (PCA) using common SNPs that were consistently genotyped in all datasets. Additionally, we included the publicly available 1000 Genomes Project dataset as a validation for the clustering of the populations. In the PCA, the AMP-PD cohort clustered together with the Fox Insight cohort and the European samples of the 1000 Genomes Project, as both consist of patients of mainly European/White descent (Figure 4). In the study that constructed the MGS that we used, the dataset consisted of participants of European ancestry,^9^ which could explain why we had the strongest correlation in *LRRK2*-PD patients with European/White ethnicity. However, the frequency of LRRK2 p.G2019S is higher in Ashkenazi Jewish and Tunisian Arab-Berber populations.^39^ This highlights the importance of deriving an MGS from these two founder populations, as it would be pertinent to further understanding the MGS effect. Combined international efforts will be required to generate, evaluate and estimate an MGS in diverse populations. The lack of diverse cohorts in large-scale genetic studies is a well-known problem,^40,41^ but more diversity is essential to overcome such limitations of polygenic scores.

**Figure 4.**
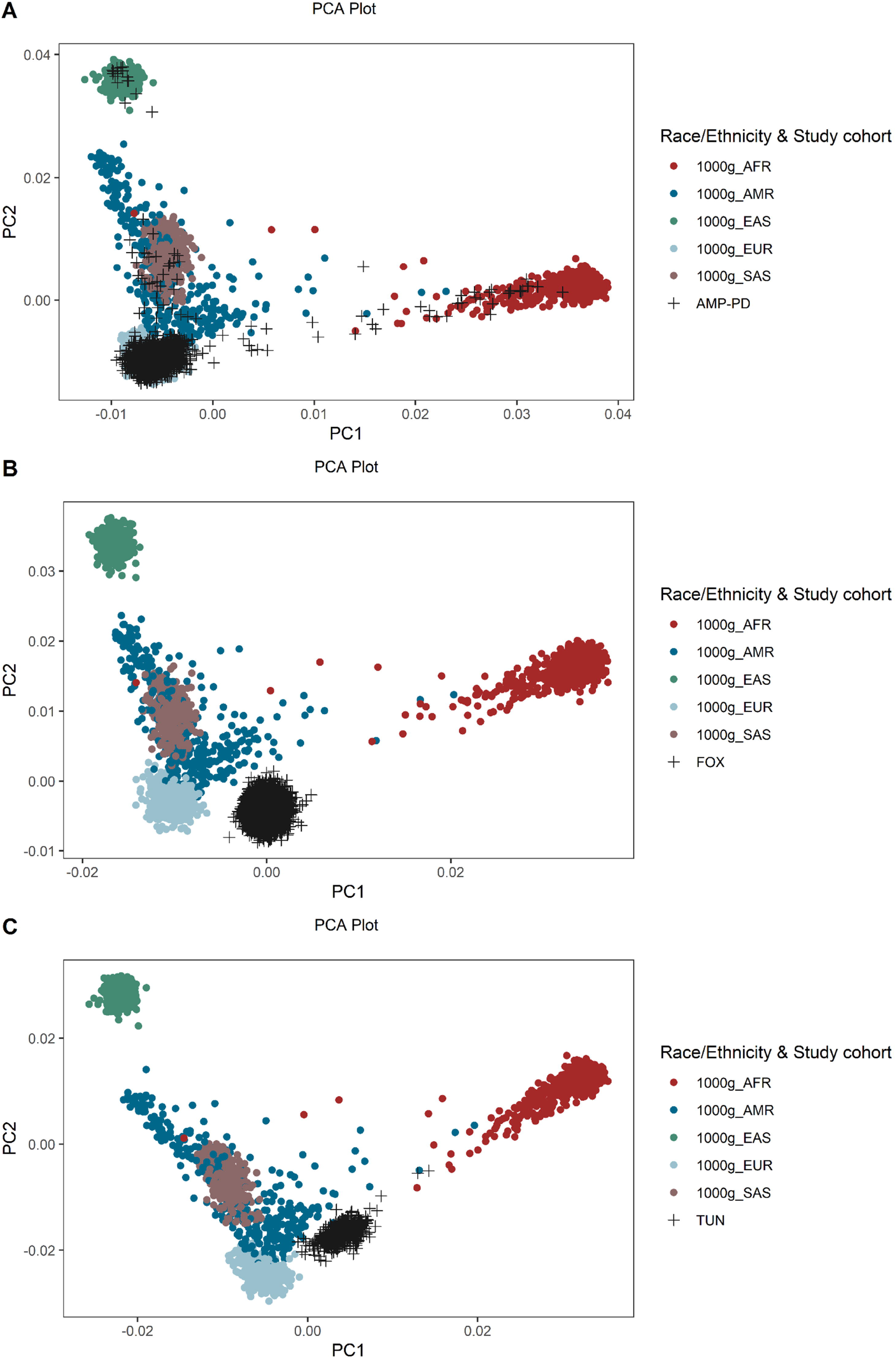
Principal component analysis (PCA) The PCA plot displays the clustering of samples included in this study. The PCA was derived from common SNPs overlapping in all data sets. The samples were colored by their study cohort and their ethnicity/race. In addition to our study cohorts (i.e., AMP-PD (**A**), Fox Insight (**B**) and Tunisian cohort (**C**)), we used the publicly available 1000 Genomes Project dataset for validation. 1000 Genomes Project super populations: AFR=African, EAS=East Asian, AMR=Ad Mixed American, EUR=European and SAS=South Asian

Limitations of our study include potential bias that comes from different data reported in the three cohorts. In terms of genetics, genotyping data were either obtained from arrays (Fox Insight and Tunisian cohort) or from WGS (AMP-PD) that could contribute to batch effects. Thus we adjusted for the cohort as a covariate in the regression model. Another limitation of the genotyping data is that we used a subset (9 out of 14) of the SNPs that constitute the MGS that were consistently present in all three datasets. However, when we just included the Fox Insight and AMP-PD dataset (*N*=281 *LRRK2*-PD patients), we could use 12 out of the 14 SNPs, and when we only included the AMP-PD dataset (*N*=127 *LRRK2*-PD patients), we could use all 14 MGS SNPs. The negative association between MGS and AAO in *LRRK2*-PD remained in these analyses as well. The main environmental/lifestyle questionnaire used in our study is the validated PD-RFQ. However, the PD-RFQ was only available from the Fox Insight and Tunisian cohort. In order to harmonize the data as much as possible, AMP-PD was not included in our environment/lifestyle analyses. The PD-RFQ, though validated, also has its own caveats. For example, pesticide exposure in a non-work setting includes any exposure to chemicals utilized to kill insects, other pests, plants, weeds, mold or mildew used in the house, garden, or on pests, which leads to an inflation of individual exposure. Diverse cultural preferences also exist that may not be captured by the lifestyle questionnaires: one example is the main source of caffeine intake (i.e., coffee, tea or soda), which varies significantly in different countries.^42^ To overcome this caveat, we stratified our data for ethnicity/race and study cohort and performed interaction analyses only on iPD patients of the Fox Insight cohort that were all of European/White ancestry. Still, caffeinated soda consumption and pesticide exposure showed an interaction with MGS in predicting AAO (Supplementary Table 6).

The association between MGS and AAO was specific to *LRRK2*-PD when the data were not stratified for any lifestyle/environmental factor. One explanation is that *LRRK2*-PD is more homogeneous compared to the patients with iPD. The causes of the disease in iPD patients can be much more diverse and this heterogeneity may overshadow the subtle effect of the MGS, which may only be valid for certain subtypes of iPD. Another potential explanation is that mitochondrial biological implications are strongly related to disease onset in *LRRK2*-PD but not in iPD. Mitochondrial abnormalities are involved in the pathogenesis of *LRRK2*-PD, such as reduced NADH dehydrogenase activity, increased mitochondrial mass, mtDNA copy number, and nuclear factor erythroid 2-related factor 2 (Nrf2) expression.^11^ Thus, an additional mitochondrial burden, reflected in a higher MGS, could lead to an earlier AAO in patients with *LRRK2*-PD. Recently, a study has shown a significant association between MGS and AAO in patients with iPD.^43^ However, a larger and different set of SNPs was used to calculate MGS. There was also no comparison of iPD with *LRRK2*-PD or other monogenic forms, where an even larger effect may be evident, as seen in our study.

Mitochondrial function can be affected by pesticide exposure, tobacco use or caffeine consumption.^10,22,23,44^ We, among others, have reported that caffeinated soda intake was associated with earlier AAO^5^ or increased PD risk.^45^ Hence, caffeinated soda appears to be different from other caffeinated beverages and potentially caffeine-independent mechanisms are driving these effects. Exposure to particular pesticides (e.g., paraquat or rotenone) has been a widely reported risk factor for PD.^10,14,16^ We observed an association between pesticide exposure and AAO in iPD (Supplementary Table 3). For patients with *LRRK2*-PD, there was an interaction between MGS and coffee consumption as well as caffeinated soda consumption. The median AAO was ∼six years and ∼11 years earlier in patients with a high MGS that consumed coffee or caffeinated soda, respectively. Additionally, the median AAO of caffeinated soda consumers with a high MGS was ∼2 years earlier in iPD patients. That is important to highlight, as there was no difference in AAO in iPD patients that did not consume caffeinated soda. Therefore, our data support a gene-environment interaction between caffeine intake and MGS. Caffeine consumption is reported as a protective factor in PD, with the exception of caffeinated soda, as described above. However, in rats, there is evidence that treatment with caffeine induces mitochondrial dysfunction in the neonatal brain.^46^

In addition to caffeine, pesticide exposure interacted with the MGS in patients with iPD exclusively. Like caffeinated soda, pesticide exposure is a risk factor in PD.^10,14,16^ Pesticides like rotenone or paraquat are known to increase mitochondrial dysfunction by inducing redox cycling or binding to complex I, which both result in the production of reactive oxygen species (ROS).^47^ As the oxidative stress for mitochondria increases, the enhanced effect of MGS in PD patients exposed to pesticides can be explained.

Our results underline the importance of including lifestyle and environment when investigating genetic associations with AAO or disease risk. Gene-lifestyle or gene-environment interactions could significantly influence the association with these traits. A recent study demonstrated that GWAS analyses could be affected by gene-environment correlations across geographic regions. The genetic correlations with socioeconomic status-related traits were significantly reduced when controlling for geographic regions.^48^ Likewise, our study shows the differences between Tunisian Arab-Berbers and European/White ancestry though a more refined investigation is warranted.

In conclusion, there was an association between the MGS and earlier AAO in patients with *LRRK2*-PD but not in iPD. Furthermore, we detected gene-environment interactions in *LRRK2*-PD and iPD. Thus, lifestyle and environmental factors interact with the MGS and affect its impact on the AAO in PD (Figure 5). Our results highlight the importance of functional studies investigating the underlying molecular mechanisms leading to the interaction between MGS, caffeine consumption, and pesticide exposure.

**Figure 5.**
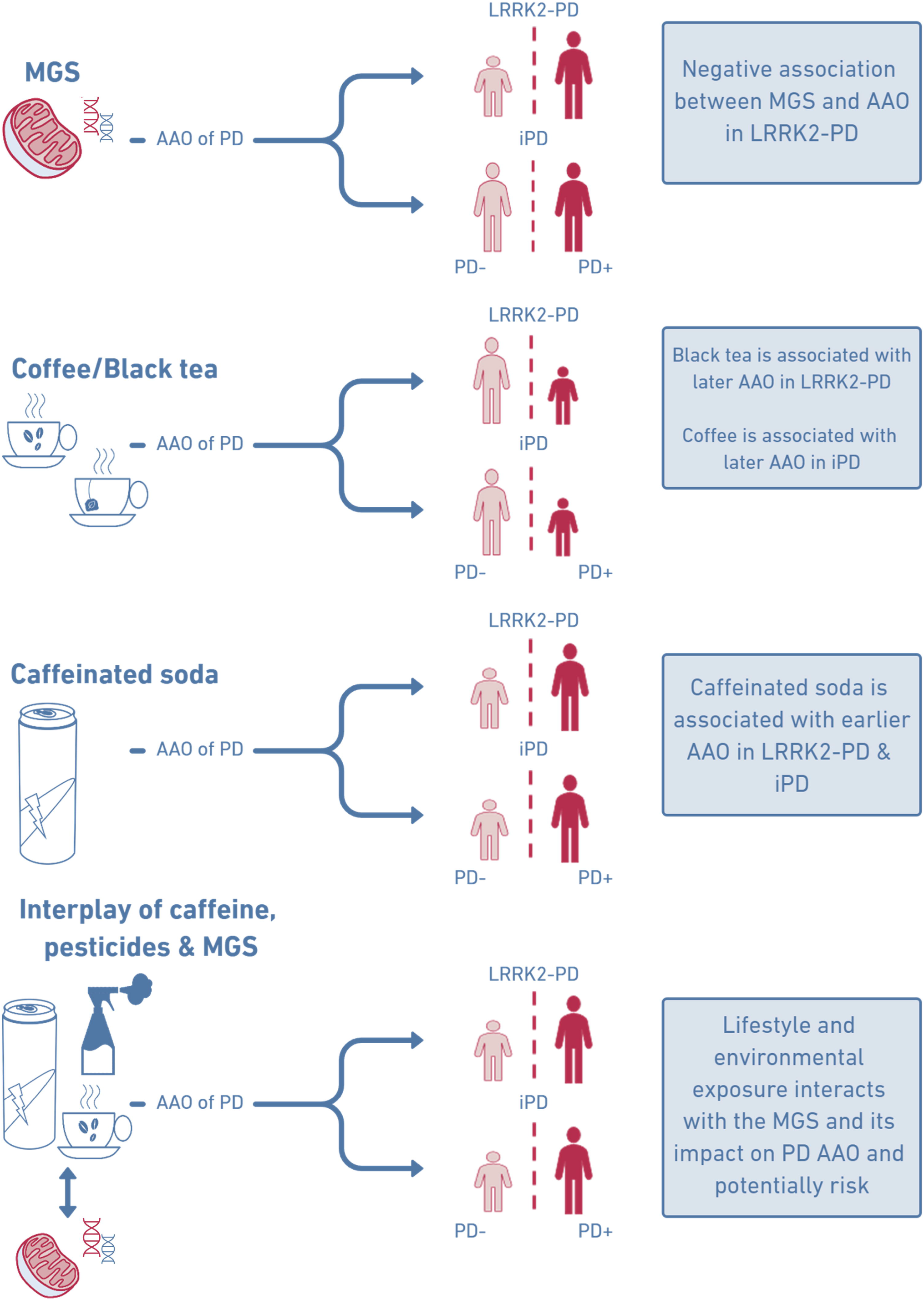
Summary of results. Caffeine consumption can be associated with later or earlier AAO in PD, depending on the beverage. Furthermore, there is evidence for gene and lifestyle/environment interactions, as in caffeine consumers and patients that were exposed to caffeine, the effect of the MGS on AAO in PD is more pronounced.

## Supporting information

STROBE statement: Reporting guidelines checklist for cohort, case-control and cross-sectional studies

## Data Availability

Data sharing is not applicable to this article as no new data were created or analysed in this study. Data used in the preparation of this manuscript were obtained from the Fox Insight database (https://foxinsight-info.michaeljfox.org/insight/explore/insight.jsp) on 18/10/2020. For up-to-date information on the study, visit https://foxinsight-info.michaeljfox.org/insight/explore/insight.jsp. Data used in the preparation of this article were obtained from the Accelerating Medicine Partnership (AMP) Parkinson's Disease (AMP PD) Knowledge Platform. For up-to-date information on the study, visit https://www.amp-pd.org.

## Acknowledgements

A special thanks to the families and patients who participated in this study.

The Fox Insight Study (FI) is funded by The Michael J. Fox Foundation for Parkinson’s Research. We would like to thank the Parkinson’s community and 23andMe research participants and employees for making this research possible.

The AMP® PD program is a public-private partnership managed by the Foundation for the National Institutes of Health and funded by the National Institute of Neurological Disorders and Stroke (NINDS) in partnership with the Aligning Science Across Parkinson’s (ASAP) initiative; Celgene Corporation, a subsidiary of Bristol-Myers Squibb Company; GlaxoSmithKline plc (GSK); The Michael J. Fox Foundation for Parkinson’s Research ; Pfizer Inc.; Sanofi US Services Inc.; and Verily Life Sciences. ACCELERATING MEDICINES PARTNERSHIP and AMP are registered service marks of the U.S. Department of Health and Human Services.

Clinical data and biosamples used in preparation of this article were obtained from the (i) Michael J. Fox Foundation for Parkinson’s Research (MJFF) and National Institutes of Neurological Disorders and Stroke (NINDS) BioFIND study, (ii) Harvard Biomarkers Study (HBS), (iii) National Institute on Aging (NIA) International Lewy Body Dementia Genetics Consortium Genome Sequencing in Lewy Body Dementia Case-control Cohort (LBD), (iv) MJFF LRRK2 Cohort Consortium (LCC), (v) NINDS Parkinson’s Disease Biomarkers Program (PDBP), (vi) MJFF Parkinson’s Progression Markers Initiative (PPMI), and (vii) NINDS Study of Isradipine as a Disease-modifying Agent in Subjects With Early Parkinson Disease, Phase 3 (STEADY-PD3) and (viii) the NINDS Study of Urate Elevation in Parkinson’s Disease, Phase 3 (SURE-PD3).

BioFIND is sponsored by The Michael J. Fox Foundation for Parkinson’s Research (MJFF) with support from the National Institute for Neurological Disorders and Stroke (NINDS). The BioFIND Investigators have not participated in reviewing the data analysis or content of the manuscript. For up-to-date information on the study, visit michaeljfox.org/news/biofind. Genome sequence data for the Lewy body dementia case-control cohort were generated at the Intramural Research Program of the U.S. National Institutes of Health. The study was supported in part by the National Institute on Aging (program #: 1ZIAAG000935) and the National Institute of Neurological Disorders and Stroke (program #: 1ZIANS003154).

The Harvard Biomarker Study (HBS) is a collaboration of HBS investigators [full list of HBS investigators found at https://www.bwhparkinsoncenter.org/biobank/ and funded through philanthropy and NIH and Non-NIH funding sources. The HBS Investigators have not participated in reviewing the data analysis or content of the manuscript.

PPMI is sponsored by The Michael J. Fox Foundation for Parkinson’s Research and supported by a consortium of scientific partners: [list the full names of all of the PPMI funding partners found at https://www.ppmi-info.org/about-ppmi/who-we-are/study-sponsors]. The PPMI investigators have not participated in reviewing the data analysis or content of the manuscript. For up-to-date information on the study, visit www.ppmi-info.org.

The Parkinson’s Disease Biomarker Program (PDBP) consortium is supported by the National Institute of Neurological Disorders and Stroke (NINDS) at the National Institutes of Health. A full list of PDBP investigators can be found at https://pdbp.ninds.nih.gov/policy. The PDBP investigators have not participated in reviewing the data analysis or content of the manuscript.

The Study of Isradipine as a Disease-modifying Agent in Subjects With Early Parkinson Disease, Phase 3 (STEADY-PD3) is funded by the National Institute of Neurological Disorders and Stroke (NINDS) at the National Institutes of Health with support from The Michael J. Fox Foundation and the Parkinson Study Group. For additional study information, visit https://clinicaltrials.gov/ct2/show/study/NCT02168842. The STEADY-PD3 investigators have not participated in reviewing the data analysis or content of the manuscript. The Study of Urate Elevation in Parkinson’s Disease, Phase 3 (SURE-PD3) is funded by the National Institute of Neurological Disorders and Stroke (NINDS) at the National Institutes of Health with support from The Michael J. Fox Foundation and the Parkinson Study Group. For additional study information, visit https://clinicaltrials.gov/ct2/show/NCT02642393. The SURE-PD3 investigators have not participated in reviewing the data analysis or content of the manuscript.

## References

1. Gabbert C, Konig IR, Luth T, et al. Coffee, smoking and aspirin are associated with age at onset in idiopathic Parkinson’s disease. J Neurol. Aug 2022;269(8):4195–4203. doi:10.1007/s00415-022-11041-x

2. Marras C, Canning CG, Goldman SM. Environment, lifestyle, and Parkinson’s disease: Implications for prevention in the next decade. Mov Disord. Jun 2019;34(6):801–811. doi:10.1002/mds.27720

3. Kline EM, Houser MC, Herrick MK, et al. Genetic and Environmental Factors in Parkinson’s Disease Converge on Immune Function and Inflammation. Mov Disord. Jan 2021;36(1):25–36. doi:10.1002/mds.28411

4. Dunn AR, Kaczorowski CC. Regulation of intrinsic excitability: Roles for learning and memory, aging and Alzheimer’s disease, and genetic diversity. Neurobiol Learn Mem. Oct 2019;164:107069. doi:10.1016/j.nlm.2019.107069

5. Luth T, Konig IR, Grunewald A, et al. Age at Onset of LRRK2 p.Gly2019Ser Is Related to Environmental and Lifestyle Factors. Mov Disord. Oct 2020;35(10):1854–1858. doi:10.1002/mds.28238

6. Klein C, Westenberger A. Genetics of Parkinson’s disease. Cold Spring Harb Perspect Med. Jan 2012;2(1):a008888. doi:10.1101/cshperspect.a008888

7. Nalls MA, Blauwendraat C, Vallerga CL, et al. Identification of novel risk loci, causal insights, and heritable risk for Parkinson’s disease: a meta-analysis of genome-wide association studies. Lancet Neurol. Dec 2019;18(12):1091–1102. doi:10.1016/S1474-4422(19)30320-5

8. Koch S, Laabs BH, Kasten M, et al. Validity and Prognostic Value of a Polygenic Risk Score for Parkinson’s Disease. Genes (Basel). Nov 23 2021;12(12)doi:10.3390/genes12121859

9. Billingsley KJ, Barbosa IA, Bandres-Ciga S, et al. Mitochondria function associated genes contribute to Parkinson’s Disease risk and later age at onset. NPJ Parkinsons Dis. 2019;5:8. doi:10.1038/s41531-019-0080-x

10. Grunewald A, Kumar KR, Sue CM. New insights into the complex role of mitochondria in Parkinson’s disease. Prog Neurobiol. Jun 2019;177:73–93. doi:10.1016/j.pneurobio.2018.09.003

11. Delcambre S, Ghelfi J, Ouzren N, et al. Mitochondrial Mechanisms of LRRK2 G2019S Penetrance. Front Neurol. 2020;11:881. doi:10.3389/fneur.2020.00881

12. Weindel CG, Martinez EL, Zhao X, et al. Mitochondrial ROS promotes susceptibility to infection via gasdermin D-mediated necroptosis. Cell. Jul 29 2022;doi:10.1016/j.cell.2022.06.038

13. Choi ML, Chappard A, Singh BP, et al. Pathological structural conversion of alpha-synuclein at the mitochondria induces neuronal toxicity. Nat Neurosci. Sep 2022;25(9):1134–1148. doi:10.1038/s41593-022-01140-3

14. Noyce AJ, Bestwick JP, Silveira-Moriyama L, et al. Meta-analysis of early nonmotor features and risk factors for Parkinson disease. Ann Neurol. Dec 2012;72(6):893–901. doi:10.1002/ana.23687

15. Li X, Li W, Liu G, Shen X, Tang Y. Association between cigarette smoking and Parkinson’s disease: A meta-analysis. Arch Gerontol Geriatr. Nov-Dec 2015;61(3):510–6. doi:10.1016/j.archger.2015.08.004

16. Breckenridge CB, Berry C, Chang ET, Sielken RL, Jr., Mandel JS. Association between Parkinson’s Disease and Cigarette Smoking, Rural Living, Well-Water Consumption, Farming and Pesticide Use: Systematic Review and Meta-Analysis. PLoS One. 2016;11(4):e0151841. doi:10.1371/journal.pone.0151841

17. Yahalom G, Rigbi A, Israeli-Korn S, et al. Age at Onset of Parkinson’s Disease Among Ashkenazi Jewish Patients: Contribution of Environmental Factors, LRRK2 p.G2019S and GBA p.N370S Mutations. J Parkinsons Dis. 2020;10(3):1123–1132. doi:10.3233/JPD-191829

18. San Luciano M, Tanner CM, Meng C, et al. Nonsteroidal Anti-inflammatory Use and LRRK2 Parkinson’s Disease Penetrance. Mov Disord. Oct 2020;35(10):1755–1764. doi:10.1002/mds.28189

19. Trinh J, Gustavsson EK, Vilarino-Guell C, et al. DNM3 and genetic modifiers of age of onset in LRRK2 Gly2019Ser parkinsonism: a genome-wide linkage and association study. Lancet Neurol. Nov 2016;15(12):1248–1256. doi:10.1016/S1474-4422(16)30203-4

20. Lai D, Alipanahi B, Fontanillas P, et al. Genomewide Association Studies of LRRK2 Modifiers of Parkinson’s Disease. Ann Neurol. Jul 2021;90(1):76–88. doi:10.1002/ana.26094

21. Tommasi S, Pabustan N, Li M, Chen Y, Siegmund KD, Besaratinia A. A novel role for vaping in mitochondrial gene dysregulation and inflammation fundamental to disease development. Sci Rep. Nov 23 2021;11(1):22773. doi:10.1038/s41598-021-01965-1

22. Goncalves DF, de Carvalho NR, Leite MB, et al. Caffeine and acetaminophen association: Effects on mitochondrial bioenergetics. Life Sci. Jan 15 2018;193:234–241. doi:10.1016/j.lfs.2017.10.039

23. Dragicevic N, Delic V, Cao C, et al. Caffeine increases mitochondrial function and blocks melatonin signaling to mitochondria in Alzheimer’s mice and cells. Neuropharmacology. Dec 2012;63(8):1368–79. doi:10.1016/j.neuropharm.2012.08.018

24. Iwaki H, Leonard HL, Makarious MB, et al. Accelerating Medicines Partnership: Parkinson’s Disease. Genetic Resource. Mov Disord. Aug 2021;36(8):1795–1804. doi:10.1002/mds.28549

25. Smolensky L, Amondikar N, Crawford K, et al. Fox Insight collects online, longitudinal patient-reported outcomes and genetic data on Parkinson’s disease. Sci Data. Feb 24 2020;7(1):67. doi:10.1038/s41597-020-0401-2

26. Semple SE, Dick F, Cherrie JW, Geoparkinson Study G. Exposure assessment for a population-based case-control study combining a job-exposure matrix with interview data. Scand J Work Environ Health. Jun 2004;30(3):241–8. doi:10.5271/sjweh.785

27. Purcell S, Neale B, Todd-Brown K, et al. PLINK: a tool set for whole-genome association and population-based linkage analyses. Am J Hum Genet. Sep 2007;81(3):559–75. doi:10.1086/519795

28. Das S, Forer L, Schonherr S, et al. Next-generation genotype imputation service and methods. Nat Genet. Oct 2016;48(10):1284–1287. doi:10.1038/ng.3656

29. McCarthy S, Das S, Kretzschmar W, et al. A reference panel of 64,976 haplotypes for genotype imputation. Nat Genet. Oct 2016;48(10):1279–83. doi:10.1038/ng.3643

30. O’Connell J, Gurdasani D, Delaneau O, et al. A general approach for haplotype phasing across the full spectrum of relatedness. PLoS Genet. Apr 2014;10(4):e1004234. doi:10.1371/journal.pgen.1004234

31. Howie BN, Donnelly P, Marchini J. A flexible and accurate genotype imputation method for the next generation of genome-wide association studies. PLoS Genet. Jun 2009;5(6):e1000529. doi:10.1371/journal.pgen.1000529

32. Genomes Project C, Auton A, Brooks LD, et al. A global reference for human genetic variation. Nature. Oct 1 2015;526(7571):68–74. doi:10.1038/nature15393

33. A language and environment for statistical computing. R Foundation for StatisticalComputing. 2020. https://www.R-project.org/

34. Wickham H. ggplot2: Elegant Graphics for Data Analysis. Springer-Verlag New York; 2016.

35. Trinh J, Amouri R, Duda JE, et al. Comparative study of Parkinson’s disease and leucine-rich repeat kinase 2 p.G2019S parkinsonism. Neurobiol Aging. May 2014;35(5):1125–31. doi:10.1016/j.neurobiolaging.2013.11.015

36. Duncan L, Shen H, Gelaye B, et al. Analysis of polygenic risk score usage and performance in diverse human populations. Nat Commun. Jul 25 2019;10(1):3328. doi:10.1038/s41467-019-11112-0

37. Martin AR, Kanai M, Kamatani Y, Okada Y, Neale BM, Daly MJ. Clinical use of current polygenic risk scores may exacerbate health disparities. Nat Genet. Apr 2019;51(4):584–591. doi:10.1038/s41588-019-0379-x

38. Caliebe A, Tekola-Ayele F, Darst BF, et al. Including diverse and admixed populations in genetic epidemiology research. Genet Epidemiol. Oct 2022;46(7):347–371. doi:10.1002/gepi.22492

39. Trinh J, Guella I, Farrer MJ. Disease penetrance of late-onset parkinsonism: a meta-analysis. JAMA Neurol. Dec 2014;71(12):1535–9. doi:10.1001/jamaneurol.2014.1909

40. Wojcik GL, Graff M, Nishimura KK, et al. Genetic analyses of diverse populations improves discovery for complex traits. Nature. Jun 2019;570(7762):514–518. doi:10.1038/s41586-019-1310-4

41. Sirugo G, Williams SM, Tishkoff SA. The Missing Diversity in Human Genetic Studies. Cell. May 2 2019;177(4):1080. doi:10.1016/j.cell.2019.04.032

42. Reyes CM, Cornelis MC. Caffeine in the Diet: Country-Level Consumption and Guidelines. Nutrients. Nov 15 2018;10(11)doi:10.3390/nu10111772

43. Dehestani M, Liu H, Sreelatha AAK, Schulte C, Bansal V, Gasser T. Mitochondrial and autophagy-lysosomal pathway polygenic risk scores predict Parkinson’s disease. Mol Cell Neurosci. Jun 13 2022;121:103751. doi:10.1016/j.mcn.2022.103751

44. Kanithi M, Junapudi S, Shah SI, Matta Reddy A, Ullah G, Chidipi B. Alterations of Mitochondrial Network by Cigarette Smoking and E-Cigarette Vaping. Cells. May 19 2022;11(10)doi:10.3390/cells11101688

45. C. Tanner Cm, C. Meng, K. Marder, S. Bressman, R. Saunders-Pullman, R. Alcalay, E. Tolosa, A. Brice, S. Goldman, B. Schuele, A. Lang, S. Goldwurm, G. Riboldazzi, J. Ferreira, C. Klein, D. Berg, K. Brockmann, M. Tazir, J. Aasly, J. Marti-Masso, J. Marti-Masso, R. Munhoz, C. Rieder, M. San Luciano, G. Mellick, C. Sue, K. Hasegawa, E. Tan, J. Langston, M. LRRK2 Cohort-Consortium (San Francisco, CA, USA). Caffeinated Drinks, LRRK2 Genotype and PD. xpresented at: International Congress of Parkinson’s Disease and Movement Disorders; 2017; Session Parkinson’s Disease: Genetics.

46. Kasala S, Briyal S, Prazad P, et al. Exposure to Morphine and Caffeine Induces Apoptosis and Mitochondrial Dysfunction in a Neonatal Rat Brain. Front Pediatr. 2020;8:593. doi:10.3389/fped.2020.00593

47. Nistico R, Mehdawy B, Piccirilli S, Mercuri N. Paraquat- and rotenone-induced models of Parkinson’s disease. Int J Immunopathol Pharmacol. Apr-Jun 2011;24(2):313–22. doi:10.1177/039463201102400205

48. Abdellaoui A, Dolan CV, Verweij KJH, Nivard MG. Gene-environment correlations across geographic regions affect genome-wide association studies. Nat Genet. Sep 2022;54(9):1345–1354. doi:10.1038/s41588-022-01158-0

