## Supplementary Table 1 for "Interaction of mitochondrial polygenic score and environmental factors in LRRK2 p.Gly2019Ser parkinsonism"

**Supplementary Table 1.** Correlation analysis between mitochondrial polygenic score and the age at onset in patients with *LRRK2*-PD and idiopathic PD, stratified by sex, study site or ethnicity.

| | **LRRK2-PD** | | **iPD** | |
|---|---|---|---|---|
| | **r** | **p** | **r** | **p** |
| **Stratified by sex** | | | **Stratified by sex** | |
| Male | -0.16 | 0.011 | -0.007 | 0.623 |
| Female | -0.16 | 0.015 | 0.009 | 0.586 |
| **Stratified by cohort** | | | **Stratified by study site** | |
| AMP-PD | -0.15 | 0.093 | 0.01 | 0.529 |
| Fox Insight | -0.15 | 0.073 | 0.002 | 0.884 |
| Tunisian cohort | -0.06 | 0.431 | 0.01 | 0.851 |
| **Stratified by ethnicity** | | | **Stratified by ethnicity** | |
| European/white | -0.16 | 0.0095 | -0.004 | 0.723 |
| Tunisian/Arab | -0.05 | 0.437 | 0.01 | 0.851 |

iPD=idiopathic Parkinson's disease, *LRRK2*-PD= Patients with PD that carry the LRRK2 G2019S variant, r = Spearman's rank correlation coefficient, p = Spearman's exploratory p-value.
