## Supplementary Table 2 for "Interaction of mitochondrial polygenic score and environmental factors in LRRK2 p.Gly2019Ser parkinsonism"

**Supplementary Table 2.** Association between lifestyle factors and the age at onset in patients with *LRRK2*-PD and idiopathic PD.

| | ***LRRK2*-PD** | | | **iPD** | | |
|---|---|---|---|---|---|---|
| | **Estimate** | **Std. Error** | **p-value** | **Estimate** | **Std. Error** | **p-value** |
| **Smoking (N=144)[1]** | | | | **Smoking (N=1759)[1]** | | |
| Lifestyle factor | 2.67 | 2.05 | 0.194 | 1.45 | 0.46 | 0.001* |
| Sex Male | 2.23 | 1.99 | 0.26 | 1.13 | 0.45 | 0.012* |
| Study Tunisian cohort | -8.65 | 2.13 | $8.2 \times 10^{-5}$* | -14.47 | 1.88 | $2.5 \times 10^{-14}$* |
| **Coffee consumption (N=136)[1]** | | | | **Coffee consumption (N=1676)[1]** | | |
| Lifestyle factor | 0.64 | 2.02 | 0.75 | 2.06 | 0.55 | 0.0002* |
| Sex Male | 3.26 | 1.92 | 0.09 | 0.64 | 0.46 | 0.16 |
| Study Tunisian cohort | -7.01 | 2.39 | 0.004* | -13.85 | 1.89 | $3.7 \times 10^{-13}$* |
| **Black tea consumption (N=130)[1]** | | | | **Black tea consumption (N=1437)[1]** | | |
| Lifestyle factor | 6.10 | 1.97 | 0.002 * | 0.71 | 0.51 | 0.162 |
| Sex Male | 3.29 | 1.89 | 0.08 | 0.74 | 0.50 | 0.14 |
| Study Tunisian cohort | -8.96 | 2.42 | 0.0003* | -14.23 | 1.88 | $6.3 \times 10^{-14}$* |
| **Green tea consumption (N=134)[1]** | | | | **Green tea consumption (N=1382)[1]** | | |
| Lifestyle factor | -2.74 | 2.20 | 0.22 | 0.87 | 0.67 | 0.192 |
| Sex Male | 2.96 | 1.90 | 0.12 | 0.56 | 0.50 | 0.27 |
| Study Tunisian cohort | -6.61 | 2.36 | 0.006* | -14.27 | 1.88 | $5.1 \times 10^{-14}$* |
| **Caffeinated soda consumption (N=133)[1]** | | | | **Caffeinated soda consumption (N=1370)[1]** | | |
| Lifestyle factor | -4.29 | 2.01 | 0.035* | -2.41 | 0.54 | $8.8 \times 10^{-6}$ * |
| Sex Male | 2.94 | 1.90 | 0.12 | 0.48 | 0.50 | 0.34 |
| Study Tunisian cohort | -8.11 | 2.38 | 0.0009* | -15.35 | 1.94 | $5.4 \times 10^{-15}$* |

iPD=idiopathic Parkinson's disease, LRRK2-PD= Patients with PD that carry the LRRK2 G2019S variant,
* p-value < 0.05
[1]glm(formula = AAO ~ Lifestyle factor + Sex + Study cohort, family = gaussian)
Baseline categories: Sex=Female and Study=Fox Insight
