## Supplementary Table 3 for "Interaction of mitochondrial polygenic score and environmental factors in LRRK2 p.Gly2019Ser parkinsonism"

**Supplementary Table 3.** Association between pesticide exposure and the age at onset in patients with LRRK2-PD and idiopathic PD.

| | ***LRRK2*-PD** | | | **iPD** | | |
|---|---|---|---|---|---|---|
| | **Estimate** | **Std. Error** | **p-value** | **Estimate** | **Std. Error** | **p-value** |
| **Pesticide exposure in a work setting (N=126)[1]** | | | | **Pesticide exposure in a work setting (N=967)[1]** | | |
| Exposure | 0.32 | 2.66 | 0.90 | -1.52 | 0.76 | 0.048 * |
| Sex Male | 3.24 | 2.02 | 0.11 | 1.18 | 0.62 | 0.06 |
| Study Tunisian cohort | -6.21 | 2.81 | 0.029* | -14.59 | 1.98 | $3.3\times10^{-13}$* |
| **Pesticide exposure in a non-work setting (N=111)[1]** | | | | **Pesticide exposure in a non-work setting (N=931)[1]** | | |
| Exposure | -1.71 | 2.23 | 0.44 | 0.51 | 1.02 | 0.617 |
| Sex Male | 3.56 | 2.12 | 0.10 | 0.74 | 0.62 | 0.23 |
| Study Tunisian cohort | -7.94 | 3.00 | 0.009* | -14.60 | 2.01 | $7.1\times10^{-13}$* |

iPD=idiopathic Parkinson's disease, *LRRK2*-PD= Patients with PD that carry the LRRK2 G2019S variant
*p-value < 0.05
[1]glm(formula = AAO ~ Exposure + Sex + Study cohort, family = gaussian)
Baseline categories: Sex=Female and Study=Fox Insight
