## Supplementary Table 4 for "Interaction of mitochondrial polygenic score and environmental factors in LRRK2 p.Gly2019Ser parkinsonism"

**Supplementary Table 4.** Interaction between the mitochondrial polygenic score and caffeine dosage on the age at onset in patients with *LRRK2*-PD and idiopathic PD

| Caffeine dosage | | | | | | |
|---|---|---|---|---|---|---|
| | ***LRRK2*-PD[1]** | | | **iPD[1]** | | |
| | **Estimate** | **Std. Error** | **p-value** | **Estimate** | **Std. Error** | **p-value** |
| **Dosage of coffee consumption (N=122)** | | | | **Dosage of coffee consumption (N=1372)** | | |
| MGS:Lifestyle factor dosage | -0.014 | 0.02 | 0.367 | -0.004 | 0.005 | 0.402 |
| Sex Male | 2.28 | 1.98 | 0.25 | 0.70 | 0.51 | 0.17 |
| Study Tunisian cohort | -9.05 | 3.47 | 0.010* | -14.04 | 1.92 | $4.7\times10^{-13}$* |
| **Dosage of caffeinated soda consumption (N=130)** | | | | **Dosage of caffeinated soda consumption (N=1148)** | | |
| MGS:Lifestyle factor dosage | -0.02 | 0.03 | 0.396 | -0.01 | 0.007 | 0.139 |
| Sex Male | 3.17 | 1.93 | 0.10 | 0.47 | 0.54 | 0.38 |
| Study Tunisian cohort | -9.95 | 3.07 | 0.002* | -14.52 | 2.02 | $1.3\times10^{-12}$* |
| **Added dosage of coffee & caffeinated soda consumption (N=134)** | | | | **Added dosage of coffee & caffeinated soda consumption (N=1414)** | | |
| MGS:Lifestyle factor dosage | -0.02 | 0.01 | 0.069 | -0.006 | 0.004 | 0.089 |
| Sex Male | 3.62 | 1.90 | 0.06 | 0.80 | 0.50 | 0.11 |
| Study Tunisian cohort | -8.49 | 3.25 | 0.010* | -14.53 | 1.92 | $6.7\times10^{-14}$* |

iPD=idiopathic Parkinson's disease, *LRRK2*-PD= Patients with PD that carry the LRRK2 G2019S variant, $MGS_{std}$=Mitochondrial polygenic score, * p-value < 0.05
[1]glm(formula = AAO ~ $MGS_{std}$ * Lifestyle factor dosage + Sex + Study cohort, family = gaussian)
