## Supplementary Table 5 for "Interaction of mitochondrial polygenic score and environmental factors in LRRK2 p.Gly2019Ser parkinsonism"

**Supplementary Table 5.** Interaction between the mitochondrial polygenic score, caffeine consumption duration or pesticide exposure duration on the age at onset in patients with *LRRK2*-PD and idiopathic PD

| Caffeine dosage | | | | | | |
|---|---|---|---|---|---|---|
| | ***LRRK2*-PD**[1] | | | **iPD**[1] | | |
| | **Estimate** | **Std. Error** | **p-value** | **Estimate** | **Std. Error** | **p-value** |
| **Duration of coffee consumption until AAO (N=48)** | | | | **Duration of coffee consumption until AAO (N=715)** | | |
| MGS:Lifestyle factor Duration | 0.003 | 0.007 | 0.72 | 0.005 | 0.004 | 0.23 |
| Sex Male | 1.14 | 2.70 | 0.68 | -0.59 | 0.55 | 0.29 |
| Study Tunisian cohort | -3.63 | 4.35 | 0.41 | -9.80 | 2.70 | 0.0003* |
| **Duration of caffeinated soda consumption until AAO (N=24)** | | | | **Duration of caffeinated soda consumption until AAO (N=354)** | | |
| MGS:Lifestyle factor Duration | -0.01 | 0.02 | 0.30 | 0.004 | 0.006 | 0.52 |
| Sex Male | 0.77 | 4.16 | 0.86 | 0.57 | 0.90 | 0.52 |
| Study Tunisian cohort | -6.02 | 6.28 | 0.35 | -20.35 | 3.80 | $1.6\times10^{-7}$* |

| Pesticide exposure duration | | | |
|---|---|---|---|
| **Duration of pesticide exposure until AAO (iPD N=153)**[2] | | | |
| | **Estimate** | **Std. Error** | **p-value** |
| MGS:Exposure duration | 0.01 | 0.02 | 0.68 |
| Sex Male | 2.26 | 1.78 | 0.21 |

iPD=idiopathic Parkinson's disease, *LRRK2*-PD= Patients with PD that carry the LRRK2 G2019S variant, $MGS_{std}$=Mitochondrial polygenic score, * p-value < 0.05

[1]glm(formula = AAO ~ $MGS_{std}$ * Lifestyle factor duration + Sex + Study cohort, family = gaussian)

[2]glm(formula = AAO ~ $MGS_{std}$ * Exposure duration + Sex, family = gaussian), only data from Fox Insight available

Baseline categories: Sex=Female and Study=Fox Insight
