## Supplementary Table 6 for "Interaction of mitochondrial polygenic score and environmental factors in LRRK2 p.Gly2019Ser parkinsonism"

**Supplementary Table 6.** Interaction between the mitochondrial polygenic score, lifestyle factors and pesticide exposure and the age at onset in patients with idiopathic PD from the Fox Insight cohort exclusively.

| | **iPD[1]** | | |
|---|---|---|---|
| | **Estimate** | **Std. Error** | **p-value** |
| **Smoking (N=1734)** | | | |
| MGS:Lifestyle factor | -0.10 | 0.09 | 0.27 |
| Lifestyle | 4.41 | 2.81 | 0.12 |
| MGS | -0.03 | 0.06 | 0.57 |
| Sex Male | 1.20 | 0.45 | 0.007* |
| **Coffee consumption (N=1651)** | | | |
| MGS:Lifestyle factor | -0.10 | 0.12 | 0.39 |
| Lifestyle | 5.13 | 3.50 | 0.14 |
| MGS | -0.006 | 0.11 | 0.96 |
| Sex Male | 0.69 | 0.46 | 0.13 |
| **Black tea consumption (N=1412)** | | | |
| MGS:Lifestyle factor | -0.07 | 0.10 | 0.10 |
| Lifestyle | 2.75 | 3.07 | 0.37 |
| MGS | -0.07 | 0.07 | 0.26 |
| Sex Male | 0.80 | 0.49 | 0.10 |
| **Green tea consumption (N=1357)** | | | |
| MGS:Lifestyle factor | -0.25 | 0.14 | 0.07 |
| Lifestyle | 8.40 | 4.15 | 0.043* |
| MGS | -0.06 | 0.06 | 0.26 |
| Sex Male | 0.66 | 0.50 | 0.19 |
| **Caffeinated soda consumption (N=1347)** | | | |
| MGS:Lifestyle factor | -0.26 | 0.11 | 0.017* |
| Lifestyle | 5.37 | 3.25 | 0.10 |
| MGS | 0.06 | 0.09 | 0.50 |
| Sex Male | 0.51 | 0.49 | 0.30 |
| **Pesticide exposure in a work setting (N=943)** | | | |
| MGS:Exposure | -0.39 | 0.16 | 0.012* |
| Exposure | 9.78 | 4.63 | 0.035* |
| MGS | -0.03 | 0.07 | 0.72 |
| Sex Male | 1.24 | 0.61 | 0.044* |
| **Pesticide exposure in a non-work setting (N=906)** | | | |
| MGS:Exposure | -0.16 | 0.21 | 0.43 |
| Exposure | 6.13 | 6.29 | 0.33 |
| MGS | 0.05 | 0.20 | 0.80 |
| Sex Male | 0.82 | 0.61 | 0.18 |

iPD=idiopathic Parkinson's disease, $MGS_{std}$=Mitochondrial polygenic score, * p-value < 0.05
[1]glm(formula = AAO ~ $MGS_{std}$ * Lifestyle factor/Exposure + Sex, family = gaussian)
Baseline categories: Sex=Female
